# Overweight/obesity as the potentially most important lifestyle factor associated with signs of pneumonia in COVID-19

**DOI:** 10.1101/2020.07.23.20161042

**Authors:** Vanessa Sacco, Barbara Rauch, Christina Gar, Stefanie Haschka, Anne L. Potzel, Stefanie Kern-Matschilles, Friederike Banning, Irina Benz, Mandy Meisel, Jochen Seissler, Andreas Lechner

## Abstract

**Objective:** The occurrence of pneumonia separates severe cases of COVID-19 from the majority of cases with mild disease. However, the factors determining whether or not pneumonia develops remain to be fully uncovered. We therefore explored the associations of several lifestyle factors with signs of pneumonia in COVID-19.

**Methods:** Between May and July 2020, we conducted an online survey of 201 adults in Germany who had recently gone through COVID-19, predominantly as outpatients. Of these, 165 had a PCR-based diagnosis and 36 had a retrospective diagnosis by antibody testing. The survey covered demographic information, eight lifestyle factors, comorbidities and medication use. We defined the main outcome as the presence vs. the absence of signs of pneumonia, represented by dyspnea, the requirement for oxygen therapy or intubation.

**Results:** Signs of pneumonia occurred in 39 of the 165 individuals with a PCR-based diagnosis of COVID-19 (23.6%). Among the lifestyle factors examined, only overweight/obesity associated with signs of pneumonia (odds ratio 2.68 (1.29 - 5.59) p=0.008). The observed association remained significant after multivariate adjustment, with BMI as a metric variable, and also after including the antibody-positive individuals into the analysis.

**Conclusions:** This exploratory study finds an association of overweight/obesity with signs of pneumonia in COVID-19. This finding suggests that a signal proportional to body fat mass, such as the hormone leptin, impairs the body’s ability to clear SARS-CoV-2 before pneumonia develops. This hypothesis concurs with previous work and should be investigated further to possibly reduce the proportion of severe cases of COVID-19.

## Introduction

COVID-19 displays a highly variable disease severity. The spectrum ranges from asymptomatic cases to respiratory or multiorgan failure (1). Pneumonia, apparent through dyspnea or the requirement for oxygen therapy or intubation, usually separates severe cases of COVID-19 from their mild counterparts (2).

What determines whether or not pneumonia develops in the course of COVID-19 remains to be fully understood (2). Several risk factors for COVID-19 related death have already been identified, such as older age, male sex and preexisting comorbidities (3). However, patients with mild disease in an outpatient setting remain understudied and therefore factors determining the general direction of the disease remain undetermined.

Lifestyle factors, such as nutritional patterns, exercise habits and the presence of overweight or obesity, influence immune function in other viral infections and immunization responses (4–6). Different underlying mechanisms and methods to boost immune function through lifestyle change have been suggested (7, 8). Whether lifestyle factors also influence immune function and thereby disease severity in the case of COVID-19 has not yet been determined. However, this is an important question because, should a connection exist, lifestyle change could become a method to reduce the proportion of severe cases. Additionally, associations between lifestyle factors and disease severity may hint at novel underlying mechanisms.

To examine the potential associations between lifestyle factors and COVID-19 severity, we conducted an online survey among individuals who had recently gone through this infection. By this approach, we were able to collect data from patients over the whole spectrum of COVID-19 severity, including many outpatients. The study participants provided information on their disease course as well as on lifestyle factors, demographics, comorbidities and medication use before and at the time of the infection.

## Material and Methods

### Study design

This survey was part of the ongoing, prospective, online cohort study *Life&Covid*, conducted in Germany since May 2020. The main inclusion criteria for this study were a recent history of COVID-19 and an age of 18 years or older. Infection with SARS-CoV-2 had to be diagnosed by PCR from a nasopharyngeal swab or, in retrospect, by antibody testing. Additional inclusion criteria were permanent residence in Germany and online informed consent. Participants were recruited through medical information web sites, social media postings and general news media. The study was approved by the ethics committee of the medical faculty of the Ludwig-Maximilians-Universität in Munich, Germany. We used the online tool Unipark (ww2.unipark.de) for this survey, which was completed by 220 individuals between May 18^th^ and July 10^th^ 2020. Participants were asked to answer all questions for the time of onset of their SARS-CoV-2 infection and before. All infections had occurred between February and June 2020.

### Outcome variables

The first outcome variable was self-reported severity of COVID-19, derived from a question with 5 possible answers:

- grade 1 – no or mild symptoms of the disease, no fever and no dyspnea
- grade 2 – strong symptoms with fever, cough, diarrhea or loss of taste, but no dyspnea
- grade 3 – strong symptoms including dyspnea, but no oxygen therapy
- grade 4 – required oxygen therapy
- grade 5 – required intubation.

For logistic regression models, we grouped grades 1 and 2 together, as well as grades 3 to 5, to create a second, binary outcome variable coding for disease with or without signs of pneumonia.

### Study hypotheses and confounding factors considered

We evaluated 8 lifestyle factors for their association with the outcome variables, 4 potentially protective and 4 potentially detrimental factors.

#### Potentially protective lifestyle factors

a. frequent exercise (a habit of at least 150 minutes of moderate or high intensity exercise per week)
b. healthy food choices (five servings of fruit and vegetables on at least 3 days per week, regular use of fresh ingredients when cooking, meat or meat products on no more than 2 days per week)
c. sufficient sleep (self-rated)
d. frequent contact with children (living with children in the same household or regular care for children, e.g. as a nanny; surrogate for regular exposure to respiratory viruses)

Potentially detrimental lifestyle factors:

a. overweight/obesity (body mass index (BMI) ≥ 25 kg/m^2^)
b. an unfavorable fat distribution (propensity for abdominal weight gain)
c. smoking (current or quit less than 5 years prior to COVID-19)
d. regular consumption of alcohol

We also considered the following potential confounding factors:

a. male sex
b. age at or above 60 years
c. preexisting, chronic pulmonary disease
d. preexisting, chronic cardiovascular disease (coronary artery disease…)
e. presence of at least one cardiovascular risk factor (hypertension, dyslipidemia, diabetes or smoking)
f. previous cancer
g. preexisting allergy
h. concomitant psychiatric disease
i. use of ace inhibitors
j. use of RAAS inhibitors (including ace inhibitors)
k. use of statins
l. frequent respiratory tract viral infections in the 3 years prior to COVID-19 (≥2 per year)
m. a respiratory tract viral infection in the 4 weeks prior to COVID-19

### Statistical methods

We used univariate and multivariate logistic regression analyses for associations with the binary outcome variable and the Mann-Whitney-U Test for the original outcome variable with 5 grades of disease severity. Generally, the significance level was set to 5%. For the initial round of screening for associations between lifestyle factors and the binary outcome variable, we also applied the Benjamini-Hochberg procedure to limit the false discovery rate to 10%. The sample size was determined by practical reasons. All questions of the online questionnaire had to be answered. Therefore, no data were missing. Sensitivity analyses are described in table 3.

## Results

Of the 220 participants of this survey, 204 had a diagnosis of SARS-CoV-2 infection by PCR (n=166) or, retrospectively, by antibody testing (n=38). We further excluded one individual with a previous organ transplantation and two individuals with active cancer on chemo- and radiotherapy, respectively. For our primary analysis, we used the remaining cohort of PCR-tested individuals (n=165). Baseline characteristics are shown in **table 1**. The major age group were those between 40 and 59 years (45.5%) and more women than men participated (66.7%). Thirty-nine individuals (23.6%) experienced dyspnea as a sign of pneumonia. Of these, 9 participants required oxygen therapy and 3 had to be intubated.

**Table 1:** Baseline characteristics of the study cohort

| **N = 165** |  | **N (%)** |
| --- | --- | --- |
| **Age** |  |  |
| 18 – 39 years |  | 46 (27.9) |
| 40 – 59 years |  | 75 (45.5) |
| 60 – 79 years |  | 42 (25.5) |
| ≥ 80 years |  | 2 (1.2) |
| **Sex** |  |  |
| Female |  | 110 (66.7) |
| Male |  | 55 (33.3) |
| **Disease severity** (in 5 grades and *binary*) |  |  |
| No or mild symptoms | 42 (25.5) | 126 (76.4) |
| Symptomatic (fever, cough, etc.) but no dyspnea | 84 (50.9) |  |
| Symptoms including dyspnea but no oxygen therapy | 27 (16.4) | 39 (23.6) |
| Required oxygen therapy | 9 (5.5) |  |
| Required intubation | 3 (1.8) |  |
| **Inpatient treatment** |  | 19 (11.5) |
| **Potentially protective lifestyle factors** |  |  |
| Frequent exercise |  | 58 (35.2) |
| Healthy food choices |  | 44 (26.7) |
| Sufficient sleep |  | 141 (85.5) |
| Frequent contact with children (i.e. regular exposure to respiratory viruses) |  | 66 (40.0) |
| **Potentially detrimental lifestyle factors** |  |  |
| Overweight/obesity (Body-mass index $\geq 25$ kg/m<sup>2</sup>) | | 63 (38.2) |
| Unfavorable fat distribution (i.e. propensity for abdominal weight gain) |  | 78 (47.3) |
| Smoking (current or quit less than 5 years ago) |  | 22 (13.3) |
| Regular consumption of alcohol |  | 57 (34.5) |
| **Comorbidities** |  |  |
| Preexisting, chronic pulmonary disease |  | 20 (12.1) |
| Preexisting, chronic cardiovascular disease (coronary artery disease, ...) |  | 16 (9.7) |
| Presence of at least one cardiovascular risk factor (hypertension, dyslipidemia, diabetes or smoking) |  | 62 (37.6) |
| Previous cancer |  | 12 (7.3) |
| Preexisting allergy |  | 61 (37.0) |
| Concomitant psychiatric disease |  | 15 (9.1) |
| **Medication use** |  |  |
| ACE inhibitors |  | 8 (4.8) |
| RAAS inhibitors (including ACE inhibitors) |  | 22 (13.3) |
| Statins |  | 10 (6.1) |
| **Exposure to other viruses** |  |  |
| Frequent respiratory tract viral infections in the three years prior to Covid-19 |  | 52 (31.5) |
| Respiratory tract infection in the 4 weeks prior to Covid-19 |  | 24 (14.5) |

Among the 8 lifestyle factors we examined in univariate logistic regression analyses, only overweight/obesity associated significantly with symptoms of pneumonia (**table 2**). In overweight/obese individuals, these were about twice as common as in lean participants (34.9 vs. 16.7%). Additionally, oxygen treatment and intubation were more frequent with overweight/obesity (**figure 1**). The BMI as a continuous variable also associated with signs of pneumonia as did the overweight/obese BMI category in the larger study cohort, which also included the participants with a positive antibody test but no PCR (**table 3**). Among the potentially confounding factors, only the presence of pulmonary or psychiatric disease linked to symptoms of pneumonia (**table 2**). Overweight/obesity remained significantly associated with signs of pneumonia in a multivariate analysis together with these two disease groups, age and sex (**table 3**).

**Table 2:** Univariate logistic regression analyses with the dependent variable ‘signs of pneumonia’

| Independent variable | Odds ratio (95% interval) | p-value |
| --- | --- | --- |
| **Potentially protective lifestyle factors** |  |  |
| Frequent exercise | 0.78 (0.36 - 1.67) | 0.51 |
| Healthy food choices | 1.11 (0.50 - 2.44) | 0.80 |
| Sufficient sleep | 0.56 (0.22 - 1.43) | 0.23 |
| Frequent contact with children | 1.39 (0.68 - 2.86) | 0.37 |
| **Potentially detrimental lifestyle factors** |  |  |
| Overweight/obesity | 2.68 (1.29 - 5.59) | 0.008* |
| Unfavorable fat distribution | 0.94 (0.46 - 1.94) | 0.87 |
| Smoking | 0.29 (0.06 - 1.28) | 0.10 |
| Regular consumption of alcohol | 0.93 (0.44 - 2.00) | 0.86 |
| **Demographic characteristics** |  |  |
| Age ≥ 60 years | 1.53 (0.70 - 3.34) | 0.28<sup>#</sup> |
| Male sex | 1.34 (0.64 - 2.83) | 0.44<sup>#</sup> |
| **Comorbidities** |  |  |
| Preexisting, chronic pulmonary disease | 2.45 (0.92 - 6.54) | 0.07<sup>#</sup> |
| Preexisting, chronic cardiovascular disease | 1.09 (0.33 - 3.58) | 0.89 |
| Presence of at least one cardiovascular risk factor | 0.83 (0.39 - 1.73) | 0.61 |
| Previous cancer | 1.08 (0.28 - 4.22) | 0.91 |
| Preexisting allergy | 0.81 (0.38 - 1.73) | 0.59 |
| Concomitant psychiatric disease | 5.99 (1.98 - 18.18) | 0.002<sup>#</sup> |
| **Medication use** |  |  |
| ACE inhibitors | 2.02 (0.46 - 8.85) | 0.35 |
| RAAS inhibitors | 1.62 (0.61 - 4.31) | 0.33 |
| Statins | 0.34 (0.04 - 2.79) | 0.32 |
| **Exposure to other viruses** |  |  |
| Frequent respiratory tract viral infections in the three years prior to COVID-19 | 1.11 (0.52 - 2.38) | 0.80 |
| Respiratory tract infection in the 4 weeks prior to COVID-19 | 1.09 (0.40 - 2.98) | 0.87 |

\* significant with a false discovery rate of 10%
<sup>#</sup> included in multivariate adjustment (table 3)

**Table 3:** Univariate and multivariate logistic regression models with the dependent variable ‘signs of pneumonia’

| **Independent variable** | **Cohort** | **Odds ratio (95% interval)** | **p-value** |
| --- | --- | --- | --- |
| Overweight/obese vs. lean | PCR (n=165) | 2.68 (1.29 - 5.59) | 0.008 |
| BMI as metric variable | PCR (n=165) | 1.083 (1.014 - 1.155)<br>per 1 kg/m<sup>2</sup> | 0.017 |
| Overweight/obese vs. lean | PCR + AB (n=201) | 2.32 (1.16 - 4.64) | 0.017 |
| Overweight/obese vs. lean<br>with adjustment for age group, sex, pulmonary and psychiatric disease | PCR (n=165) | 2.33 (1.06 - 5.12) | 0.036 |

PCR: participants with a diagnosis of SARS-CoV-2 infection by PCR
AB: participants with a retrospective diagnosis by antibody testing

**Figure 1:**
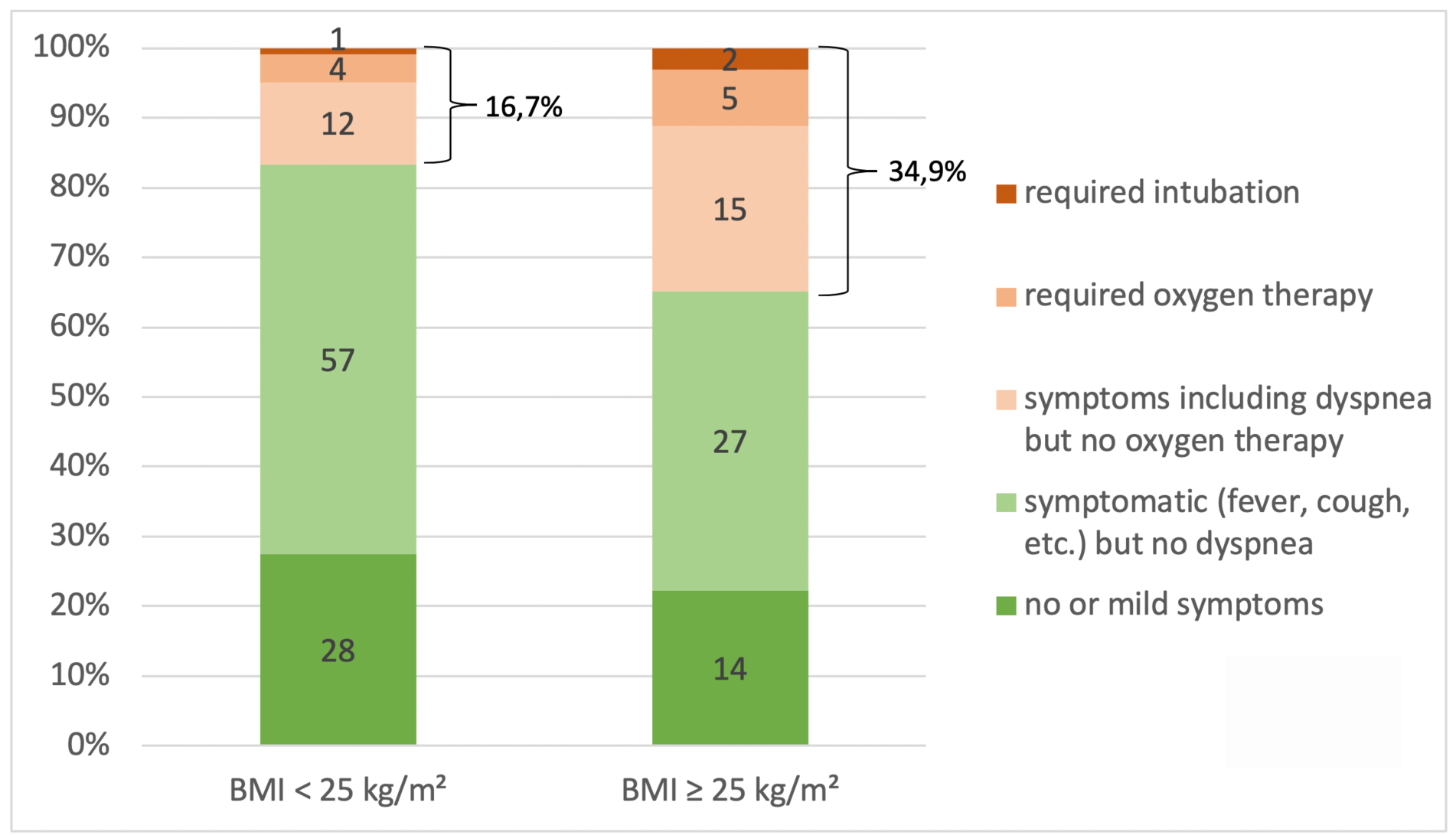
Severity of COVID-19 in lean (BMI < 25 kg/m^2^; n=102) vs. in overweight/obese individuals (BMI ≥ 25 kg/m^2^; n=63); cases with signs of pneumonia shown in shades of red; numbers = n; p for between-group difference = 0.037 (Mann-Whitney-U Test)

## Discussion

We found that overweight/obesity associated with signs of pneumonia, and thus with a higher severity of COVID-19. Other lifestyle factors, such as body fat distribution, exercise level, smoking and nutritional habits, did not show an association. Our findings extend previous work, which identified morbid obesity as a risk factor for death in COVID-19 patients (3). Our study adds that an increased risk for a higher disease severity may already start in the overweight BMI range and that excess body fat may not only influence the risk of death, but also the risk of pneumonia early on in disease development.

We had initially hypothesized that metabolic syndrome was the link between obesity and a higher COVID-19 severity, but the lack of associations with fat distribution and exercise level in our study questions this hypothesis. It rather seems that fat mass itself is the determining factor. This leads to the intriguing hypothesis that a factor proportional to fat mass negatively influences the body’s ability to clear a SARS-CoV-2 infection before pneumonia develops. A likely candidate for this would be leptin, which can, through SOCS-3, downregulate the type I interferon response (9). This response has recently been shown to be critical for timely SARS-CoV-2 clearance (9, 10).

Other lifestyle factors were unrelated to disease severity in our survey, but we would not conclude that they are irrelevant in the context of COVID-19. Rather, their effect size may be smaller and thus undetectable in our sample. Additionally, the reporting bias in survey items regarding, for example, physical activity or nutrition may be higher than in items asking for measurements such as weight and height.

This study has to be considered exploratory and hypothesis forming only, because we relied on self-reported information gathered retrospectively and because our sample is not population-based. Additionally, by design, individuals who died from COVID-19 were not included in this analysis. On the other hand, the representation of the understudied mild disease cases not treated in a hospital is a strength of this study.

Together with other work (10), our study suggests a detrimental effect of signals proportional to body fat mass, such as leptin, on the immune response to SARS-CoV-2. Should this hypothesis be proven correct, it could lead to new methods to reduce the proportion of severe cases of COVID-19. These methods could include a recommendation of weight loss, but also pharmacologic approaches to inhibit adipose tissue-derived signals, such as leptin, in infected individuals. Furthermore, overweight/obesity could serve as a risk marker to prioritize individuals for vaccination or proactive treatment of the infection.

## Data Availability

Data from this study are available from the corresponding author upon reasonable request and within the framework of European data protection laws.

## Acknowledgments

We thank all participants in the *Life&Covid* Study for their invaluable contribution.

## Author Contributions

Conceptualization, all; Formal Analysis, V.S., B.R., A.L.; Data Curation, V.S., B.R.; Writing - Original Draft, V.S., B.R., A.L.; Writing – Review & Editing, all; Visualization, V.S.; Supervision, A.L.; Project Administration, V.S.; Funding Acquisition A.L., J.S.; A.L. is the guarantor of this work. This work was not published previously.

## Data availability

Data from the *Life&Covid* study are available from the corresponding author upon reasonable request and within the framework of European data protection laws.

